# Neuroanatomical dimensions in recent-onset depression: clinical profiles, inflammatory markers, and proteomic ageing

**DOI:** 10.64898/2026.06.01.26354320

**Authors:** Paris Alexandros Lalousis, Louise Moles, Mathilde Antoniades, Wenyi Xiao, Amalie C. M. Couch, Guray Erus, Pranav Thokachichu, Dhivya Srinivasan, Yong Fan, Rachel D. Woodham, Danilo Arnone, Stephen R. Arnott, Taolin Chen, Ki Sueng Choi, Cherise Chin Fatt, Benicio N. Frey, Vibe G. Frokjaer, Melanie Ganz, Beata R. Godlewska, Stefanie Hassel, Keith Ho, Andrew M. McIntosh, Kun Qin, Susan Rotzinger, Matthew D. Sacchet, Jonathan Savitz, Haochang Shou, Aleks Stolicyn, Irina Strigo, Stephen C. Strother, Duygu Tosun, Teresa A. Victor, Dongtao Wei, Toby Wise, Roland Zahn, Ian M. Anderson, J.F. William Deakin, W. Edward Craighead, Boadie W. Dunlop, Rebecca Elliott, Qiyong Gong, Ian H. Gotlib, Catherine J. Harmer, Sidney H. Kennedy, Gitte M. Knudsen, Helen S. Mayberg, Martin P. Paulus, Jiang Qiu, Madhukar H. Trivedi, Heather C. Whalley, Chao-Gan Yan, Allan H. Young, Grace Jacobs, Nora Penzel, Raimo K.R. Salokangas, Eva Meisenzahl, Jarmo Hietala, Alessandro Bertolino, Rebekka Lencer, Stephen Wood, Christos Pantelis, Georg Romer, Dominic Dwyer, Joseph Kambeitz, Lana Kambeitz-Ilankovic, Frauke Schultze-Lutter, Stephan Ruhrmann, Nicholas M. Barnes, Rachel Upthegrove, Nikolaos Koutsouleris, Christos Davatzikos, Cynthia H.Y. Fu

**Author notes:** Joint first authors (Paris Alexandros Lalousis, Louise Moles). **Authors for correspondence:** Paris Alexandros Lalousis, PhD -, Cynthia Fu, MD, PhD –;, Christos Davatzikos, PhD –.

## Abstract

**Background:** Major depressive disorder (MDD) is clinically heterogeneous, hindering identification of reproducible biomarkers. Using a semi-supervised machine learning approach, HYDRA, we previously identified two neuroanatomical dimensions from structural MRI in medication-free MDD from COORDINATE-MDD consortium. These dimensions (D1, D2) showed differential responses to selective serotonin reuptake inhibitor (SSRI) antidepressants and placebo. External replication in UK Biobank linked D2, characterized by widespread subtle neuroanatomical reductions, to an immuno-metabolic profile. Here, we examined whether these dimensions are detectable early in the course of illness.

**Methods:** We applied the pre-trained model to structural MRI data from the multisite PRONIA cohort, comprising individuals with recent-onset depression (ROD; n = 377; mean age 25.8 years, SD 6.0; 51.3% female) and healthy controls (n = 267; mean age 25.5 years, SD 6.4; 61.0% female). Participants were assigned to clusters (C1, C2) corresponding to the previously identified dimensions (D1, D2). Clusters were compared on clinical symptom profiles, peripheral inflammatory markers, and in a subset (n = 107), proteomic ageing indices.

**Results:** Two neuroanatomical clusters were identified in PRONIA. C1 (n = 265) showed higher negative symptom severity and elevated interleukin-2 levels. C2 (n = 140) was associated with higher residual proteomic age. Overall depressive symptom severity did not differ significantly between clusters.

**Conclusions:** Neuroanatomical dimensions of MDD are reproducible and detectable at illness onset. Associations with negative symptom severity, inflammatory signalling, and proteomic ageing suggest these dimensions capture biologically meaningful heterogeneity early in depression. These findings support a biologically informed framework for stratified treatment approaches in MDD.

## Introduction

Major depressive disorder (MDD) is a leading cause of disability worldwide, affecting more than 280 million individuals each year (World Health Organisation, 2023). Core symptoms include low mood and/or loss of interest or pleasure, alongside changes in sleep, appetite, energy, and cognitive impairments (American Psychiatric Association, 2000). Despite its clinical impact, diagnosis remains based on symptom criteria rather than underlying biological mechanisms. MDD is highly clinically heterogeneous, contributing to diagnostic uncertainty and variable treatment response (Cipriani et al., 2018; Kamp et al., 2025). This categorical framework groups together individuals who may differ substantially in pathophysiology, obscuring meaningful biological signals. Such heterogeneity reflects equifinality, whereby multiple distinct biological processes can give rise to similar symptom profiles (Fu et al., 2019; Zahn, 2025; Di Biase et al., 2026).

Data-driven approaches offer a strategy to address this heterogeneity. HYDRA (Heterogeneity through Discriminant Analysis) is a semi-supervised machine learning method that simultaneously distinguishes patients from controls while partitioning patients according to disease-related variability, rather than assuming a single disease process (Varol et al., 2017). Using this approach, we previously identified two neuroanatomical dimensions in medication-free individuals with MDD from the COORDINATE-MDD consortium (n = 801 MDD, n = 765 healthy controls (HC); Fu et al., 2024). Dimension 1 (D1) was characterised by largely preserved grey and white matter volumes and showed a favourable response to selective serotonin reuptake inhibitor (SSRI) antidepressant treatment but not to placebo, whereas Dimension 2 (D2) showed subtle but widespread volumetric reductions and limited differentiation between SSRI and placebo response. External replication in the UK Biobank (n = 37,235; Xiao et al., 2025) further demonstrated that D2 was associated with cognitive impairment, childhood adversity, self-harm, a pro-atherogenic and inflammatory metabolic profile, and distinct genetic loci, suggesting a distinct immuno-metabolic subtype (Lamers et al., 2020; Xiao et al., 2025).

Peripheral inflammation has been consistently implicated in MDD, most reliably in chronic and recurrent illness and with effect sizes that are typically modest (Köhler et al., 2014; Osimo et al., 2020). Inflammatory markers have been linked to specific symptom domains, including anhedonia, fatigue, psychomotor slowing and disturbed sleep (Felger et al., 2016; Lee and Giuliani, 2019), and to poorer antidepressant response (Strawbridge et al., 2015; Arteaga-Henríquez et al., 2019). Inflammatory alterations are not solely a consequence of illness duration as elevated cytokines have been observed in first-episode and recent-onset MDD and genetic and longitudinal studies suggest a potential causal contribution to MDD risk (Pariante, 2017; Khandaker et al., 2018).

Accelerated biological ageing has also been reported in MDD across multiple modalities, including telomere shortening, epigenetic clocks and MRI-derived brain age (Schutte and Malouff, 2015; Han et al., 2018), with greater effects in chronic and severe illness. We have observed brain age acceleration in particular with the D2 dimension, which increases with chronological age (Sharma et al., 2026). Proteomic ageing clocks, which estimate biological age from circulating plasma protein expression, are robust predictors of mortality and multimorbidity at the population level (Argentieri et al., 2024; Lehallier et al., 2019). Chronic low-grade inflammation has been implicated in accelerated biological ageing, suggesting a potential mechanism through which immune dysregulation may contribute to the structural and biological alterations observed in specific subgroups of MDD (Furman et al., 2019).

The PRONIA cohort is a deeply phenotyped, multisite European sample of young adults with recent-onset depression or psychosis, providing a unique cohort to examine these questions in early illness (Koutsouleris et al., 2018). A previous PRONIA analysis identified two neuroanatomical clusters, preserved and impaired, across recent-onset depression and recent-onset psychosis that mapped to distinct clinical phenotypes, with chronic-illness samples preferentially assigned to the impaired cluster on external validation (Lalousis et al., 2022). Multivariate brain–blood signatures further characterised recent-onset depression by elevated peripheral inflammatory markers and grey matter reductions in limbic and temporal regions, distinct from the inflammatory profile in recent-onset psychosis (Popovic et al., 2026). The present analysis extends these findings by applying the externally pre-trained, MDD-specific HYDRA model derived from COORDINATE-MDD (Fu et al., 2024) without retraining to recent-onset depression alone, and by incorporating a proteomic ageing analysis (n = 107) not previously reported in this cohort.

We sought to investigate whether the neuroanatomical dimensions derived from COORDINATE-MDD are detectable early in the illness course and whether they are associated with peripheral inflammatory markers and proteomic ageing. If present at illness onset, these dimensions would more likely reflect core neurobiological vulnerability than secondary effects of illness burden. We tested three hypotheses: (i) D1 and D2 can be identified in an independent early-course sample; (ii) the dimensions are associated with distinct clinical symptom profiles; and (iii) they show different associations with peripheral inflammation and biological ageing, consistent with an immuno-metabolic mechanism contributing to MDD heterogeneity.

## Methods

### Participants

Data were drawn from the PRONIA study (Personalised Prognostic Tools for Early Psychosis Management; Koutsouleris et al., 2018), a multisite longitudinal cohort conducted across seven sites in five European countries: Germany (Munich, Cologne), Finland (Turku), Italy (Milan, Udine), Switzerland (Basel), and United Kingdom (Birmingham). The present analysis included participants with recent-onset depression (ROD) and healthy controls. ROD was defined as first lifetime major depressive episode within 3 months of a DSM-IV diagnosis of major depressive disorder (American Psychiatric Association, 2000; Koutsouleris et al., 2018). All participants were aged 15 - 40 years at study entry. Healthy controls had no current or past psychiatric disorders as assessed by structured clinical interview and were matched at the group level on key demographic variables. All participants provided written informed consent. The study was approved by the local ethics committees at each participating site.

### Structural MRI acquisition and processing

High-resolution T1-weighted structural MRI scans had been acquired at baseline across PRONIA sites with harmonised acquisition protocols. Images were visually inspected, defaced, anonymized, and pre-processed using the CAT12 toolbox (Gaser et al., 2024) for tissue segmentation, spatial normalization to MNI space, modulation, and quality control, following PRONIA procedures (Koutsouleris et al., 2018;Lalousis et al., 2022). Regional grey matter volumetric features were then extracted to match the feature space of the original HYDRA discovery study (Fu et al, 2024).

### HYDRA cluster assignment

The pre-trained HYDRA model derived from the COORDINATE-MDD discovery cohort (Fu et al., 2024) was applied to PRONIA ROD participants without retraining. HYDRA is a semi-supervised machine learning method that models disease heterogeneity by identifying multiple discriminative patterns relative to healthy controls using a set of maximum-margin hyperplanes (Varol et al., 2017).

Individual structural MRI-derived regional volumetric features were projected onto the trained model to obtain dimension expression scores for each participant. Participants were assigned to the dimension with the highest expression score (D1 or D2). Participants expressing both dimensions were classified as combined D1&D2, whereas those not showing an expression score for either dimension were classified as neither D1/D2. Group membership was used in subsequent analyses

### Clinical assessments

Baseline clinical measures included the Beck Depression Inventory (BDI-II; Beck et al., 1996), Positive and Negative Syndrome Scale (PANSS; Kay et al., 1987), and Structured Clinical Assessment for Negative Symptoms (SANS; Andreasen, 1983). Prodromal symptoms for psychosis were assessed using the Structured Interview for Prodromal Syndromes (SIPS; Miller et al., 2003). PANSS was administered across the full PRONIA cohort to enable transdiagnostic comparison; in the present analysis we report PANSS scores in the recent-onset depression sample to allow direct comparison with prior PRONIA work (Lalousis et al., 2022) and to capture symptom dimensions not fully indexed by the BDI-II.

### Inflammatory biomarkers

Peripheral blood biomarker data acquired within PRONIA were analysed, including high-sensitivity C-reactive protein (CRP), interleukin-6 (IL-6), interleukins (IL-1β, IL-1RA, IL-2, IL-4), interferon-γ (IFN-γ), tumour necrosis factor-α (TNF-α), S100B, brain-derived neurotrophic factor (BDNF), and transforming growth factor-β (TGF-β). Assays and quality control procedures were conducted according to the PRONIA protocols. The cytokine panel analysed here is broader than that reported in Lalousis et al. (2022), which did not include IL-2, IL-1β, IL-4, or IFN-γ.

### Proteomic ageing

Proteomic ageing metrics were derived from plasma proteomic data. Plasma proteins were quantified using liquid chromatography-mass spectrometry (LC-MS) with an Orbitrap mass spectrometer at the Max Planck Institute of Biochemistry in Munich. A total of 340 plasma proteins were initially quantified. After excluding proteins with more than 20% missing values, 277 proteins were retained as proteomic features for downstream analyses. Proteomic age was estimated using an established model trained to predict chronological age from circulating protein expression profiles. The proteomic age gap was calculated as the difference between estimated proteomic age and chronological age. To adjust for chronological age, residual proteomic age was computed by regressing proteomic age on chronological age and extracting residuals. Proteomic ageing measures were compared across HYDRA-defined clusters using univariate analyses. Associations between proteomic and chronological age were examined using cluster-stratified linear models.

## Statistical Analysis

Between-group differences in clinical and biological measures were assessed using independent-samples t-tests (continuous variables) and chi-squared tests (categorical variables). Multiple comparison correction was applied using the Benjamini-Hochberg false discovery rate (FDR) at q = 0.05.

## Results

### Replication of dimensions (D1, D2) in PRONIA ROD clusters (C1, C2)

The final sample comprised of 377 ROD participants (mean age 25.66 years (SD= 5.98); 51.3% female) and 267 healthy control participants (mean age 25.5 years (SD=6.4); 61% female) (Table 1).

**Table 1.** Demographic and diagnostic characteristics by cluster.

|  | C1 | C2 | Combined<br>C1+C2 | Neither<br>C1/C2 |
| --- | --- | --- | --- | --- |
| Total | 265 | 140 | 15 | 224 |
| Female | 132 | 84 | 10 | 132 |
| Recent-onset depression | 175 | 66 | 1 | 135 |
| Healthy controls | 90 | 74 | 14 | 89 |
| Age | 25.71 ± 6.21 | 26.28 ± 6.11 | 26.28 ± 6.09 | 25.93 ± 6.18 |

Total number, number female, number recent-onset depression, and number of healthy controls is presented for each cluster (C1, C2, Combined C1+C2, Neither C1/C2). Mean age is presented with standard deviation in years. C1 and C2 correspond to neuroanatomical clusters; “Combined C1+C2” indicates expression of both dimensions, and “Neither C1/C2” indicates no expression of either dimension.

Application of the pre-trained HYDRA model to the PRONIA ROD cohort identified two clusters consistent with the discovery study (Figure 1). Participants in PRONIA were assigned to clusters (C1, C2) corresponding to the previously defined dimensions (D1, D2) derived from the COORDINATE-MDD discovery sample. The preserved cluster (C1) comprised 265 participants and the impaired cluster (C2) comprised 140 participants. Within C1, 175 were ROD participants and 90 were HC and within C2, 66 were ROD and 74 were HC. The clusters did not differ significantly in age, sex, or education (all *p* > 0.05) (Table 1).

**Figure 1.**
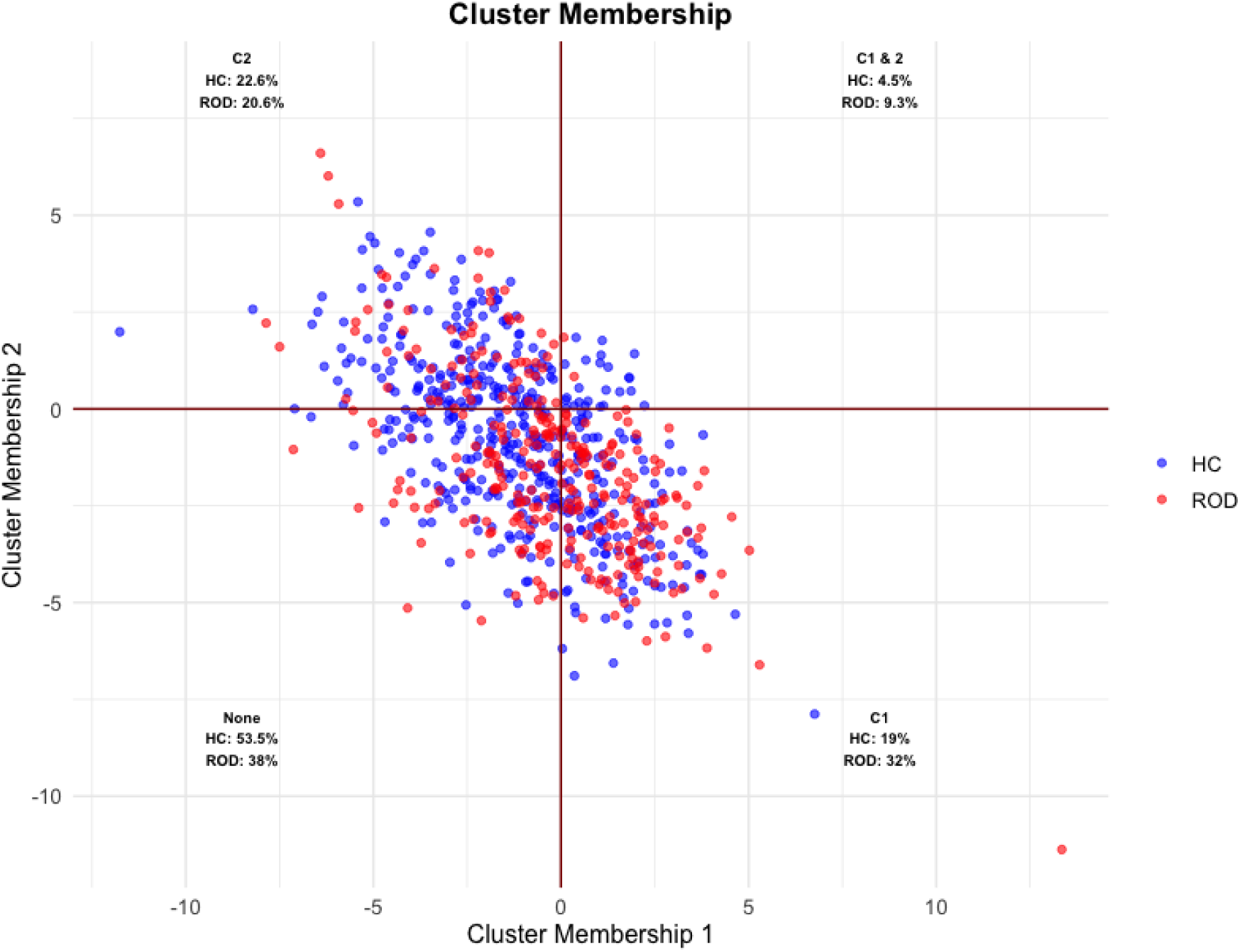
HYDRA cluster membership in the PRONIA cohort. Scatter plot of dimension expression scores for Cluster Membership 1 (C1) (x-axis) and Cluster Membership 2 (C2) (y-axis) derived from the pre-trained HYDRA model applied to PRONIA participants. Each point represents an individual participant: healthy controls (HC; blue) and recent-onset depression (ROD; red). The intersection of the two axes (at zero) defines four quadrants corresponding to cluster assignments. Bottom right: C1 (preserved; HC 19%, ROD 32%); top left: C2 (impaired; HC 22.6%, ROD 20.6%); top right: Combined C1 & C2 (combined expression; HC 4.5%, ROD 9.3%); bottom left: Neither C1/C2 (unassigned; HC 53.5%, ROD 38%). Percentages refer to the proportion of each diagnostic group assigned to that quadrant. The majority of participants cluster near the origin, with ROD participants more frequently assigned to C1 and HC more frequently unassigned.

**Figure 2.**
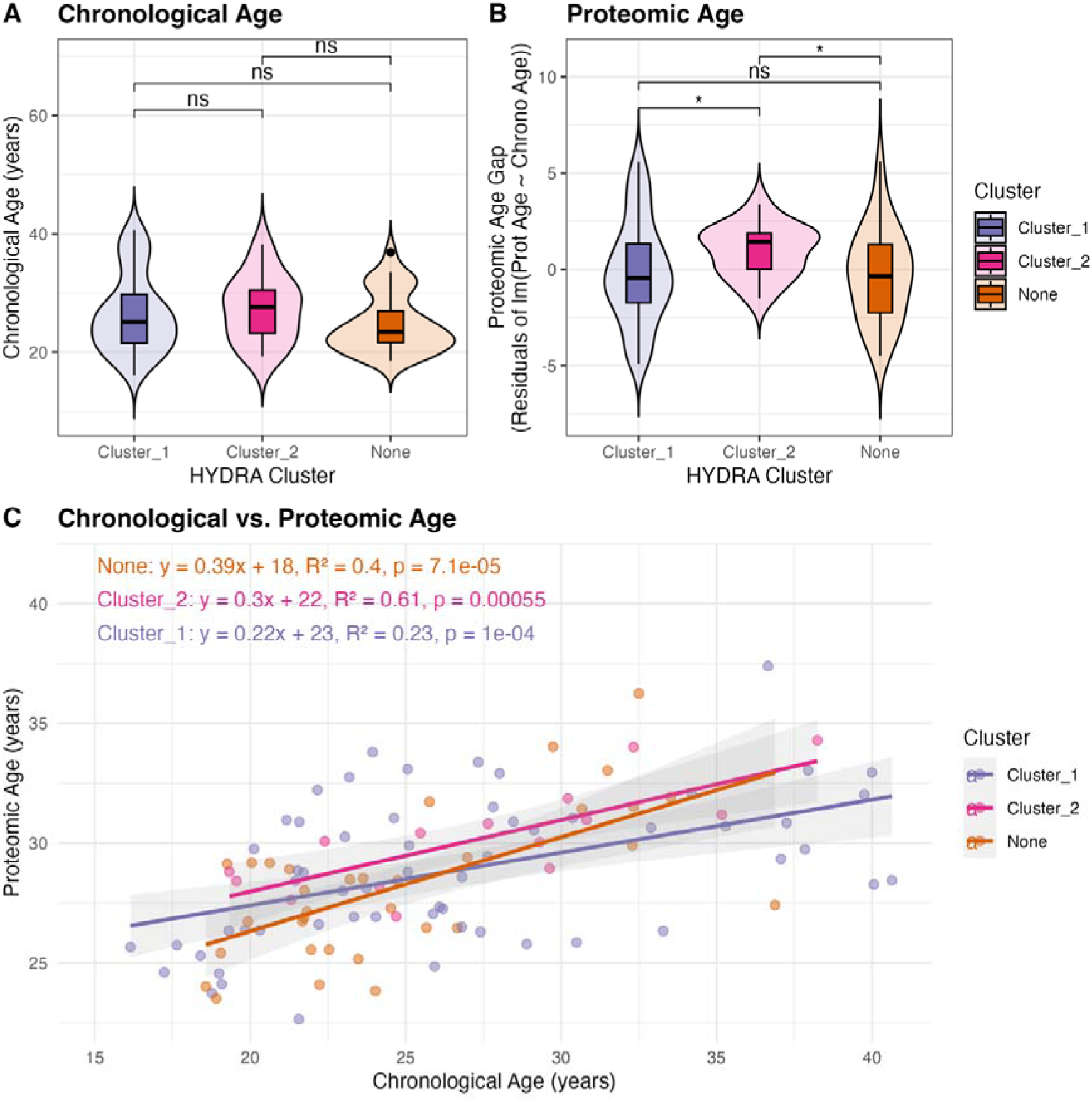
Proteomic ageing metrics by HYDRA cluster. Proteomic ageing was examined across HYDRA-derived clusters (Cluster 1/C1, preserved; Cluster 2/C2, impaired; None, unassigned) in a subset of recent-onset depression (ROD) participants with available plasma proteomic data (C1: n=59; C2: n=15; None: n=33). Chronological age did not differ between clusters (all ns), confirming groups were age-matched. Proteomic age gap (proteomic age minus chronological age) did not differ significantly between clusters (all pairwise comparisons ns). Residual proteomic age, adjusted for chronological age by linear regression, was significantly higher in C2 relative to the unassigned group (*p < 0.05), with a non-significant difference between C2 and C1 (ns) and between C1 and None (ns). Violin plots display the full data distribution with embedded box plots (median and interquartile range). Cluster-stratified linear regression of chronological versus proteomic age. The strongest association was observed in C2 (y=0.30x+22, R² = 0.61, *p* = 0.00055), followed by None (y=0.39x+18, R² = 0.40, *p* = 7.1×10⁻⁵) and C1 (y=0.22x+23, R² = 0.23, *p* = 1×10⁻⁴). Shaded regions represent 95% confidence intervals.

### Clinical features of clusters

C1 showed significantly higher negative symptom severity compared to C2, as measured by PANSS negative scores (t = 2.060, *p* = 0.041; SANS total: C1 mean = 26.879; C2 mean = 18.023; t = 2.733, p = 0.007). No significant differences were observed in PANSS positive symptoms (t = 0.628, *p* = 0.531), PANSS general psychopathology (t = 1.228, *p* = 0.222), or depressive symptom severity as measured (BDI-II; t = 1.162, *p* = 0.248) (Table 2).

**Table 2.** Baseline clinical characteristics of recent-onset depression participants by cluster.

| Measure | C1 | C2 | t | df | p |
| --- | --- | --- | --- | --- | --- |
| PANSS Positive | 7.94 | 7.77 | 0.63 | 131.3 | 0.531 |
| PANSS Negative | 13.17 | 11.33 | 2.06 | 86.5 | **0.042\*** |
| PANSS General | 28.51 | 26.91 | 1.23 | 111.1 | 0.222 |
| SANS total | 26.88 | 18.02 | 2.73 | 88.1 | **0.008\*** |
| BDI-II total | 28.07 | 25.54 | 1.16 | 98.3 | 0.248 |

Mean is presented for each cluster (C1, C2) in clinical assessments at baseline. Summary of clinical measures (mean). Values for C1 and C2 are presented as mean scores. Significant p values are shown in bold\*. PANSS, Positive and Negative Syndrome Scale; SANS, Scale for the Assessment of Negative Symptoms; BDI-II, Beck Depression Inventory-II.

### Inflammatory markers

IL-2 was significantly elevated in C1 after FDR correction ( *p* = 0.032), with uncorrected trends for IL-4 and IL-1β, and no significant differences for the remaining 8 biomarkers (Table 3).

**Table 3.** Peripheral inflammatory and neurotrophic markers in recent-onset depression participants by HYDRA cluster.

| Marker | C1 | C2 | t | df | 95% CI | p | FDR p |
| --- | --- | --- | --- | --- | --- | --- | --- |
| IFN-γ | 5.67 (18.59) | 2.09 (1.13) | 1.31 | 46.9 | -1.90 to 9.06 | 0.196 | 0.17 |
| IL-1RA | 752.45<br>(902.57) | 748.36<br>(670.33) | 0.02 | 38.2 | -419.31 to<br>427.58 | 0.984 | 0.17 |
| IL 4 | 12.40 (13.13) | 8.03 (6.37) | 1.78 | 56.6 | -0.55 to 9.31 | 0.08 | 0.1 |
| S100B | 40.05 (56.81) | 73.22 (104.73) | -1.2 | 19.5 | -88.99 to 22.66 | 0.229 | 0.17 |
| IL-1β | 0.98 (0.88) | 0.66 (0.56) | 1.72 | 45.3 | -0.06 to 0.7 | 0.093 | 0.1 |
| IL 2 | 1.67 (2.13) | 0.77 (0.82) | 2.44 | 61.7 | 0.16 to 1.64 | 0.018 | **0.032\*** |
| IL 6 | 1.07 (2.19) | 0.88 (0.95) | 0.47 | 59.7 | -0.61 to 0.97 | 0.642 | 0.35 |
| TNF-α | 1.94 (1.24) | 1.59 (1.46) | 0.87 | 24.9 | -0.47 to 1.16 | 0.392 | 0.25 |
| CRP | 2.38 (5.60) | 1.69 (2.20) | 0.71 | 61.5 | -1.26 to 2.64 | 0.482 | 0.28 |
| TGF-β | 367.95<br>(409.65) | 392.50<br>(350.71) | -0.2 | 32.9 | -236.11 to<br>187.02 | 0.815 | 0.43 |
| BDNF | 23.82 (9.89) | 21.13 (6.96) | 1.21 | 40.5 | -1.80 to 7.17 | 0.233 | 0.17 |

Means and standard deviations (SD) in parentheses are presented for each marker for C1 and C2 clusters. Statistical comparisons were performed using independent-samples t-tests. Confidence intervals represent the 95% CI for the C1–C2 mean difference. P values were corrected for multiple comparisons using the false discovery rate. Significant values are shown in bold\*. Units are pg/mL for all markers except CRP, which is reported in mg/L, and TGF-β and BDNF, which are reported in ng/mL. IFN-γ , interferon gamma; IL, interleukin; IL-1β , interleukin-1 beta; IL-1RA, interleukin-1 receptor antagonist; TNF-α , tumour necrosis factor alpha; CRP, C-reactive protein; TGF-β , transforming growth factor beta; BDNF, brain-derived neurotrophic factor.

### Proteomic ageing

Proteomics data was available in 107 ROD participants: C1 (n = 59, mean age 26.6 years, SD 6.6, 44.1% female); C2 (n = 15, mean age 27.3 years, SD 5.6, 66.7% female); neither C1/C2 (n = 33, mean age 24.8 years, SD 5.0, 48.5% female). The groups did not differ significantly in chronological age (all pairwise *p* > 0.05). Raw proteomic age gap did not differ significantly between clusters (all *p* > 0.05). However, residual proteomic age (adjusting for chronological age) was higher in C2 (impaired cluster) relative to C1 (preserved cluster) and neither C1/C2. Estimated marginal means of age-adjusted proteomic age showed a graded pattern: C2 > C1 > neither C1/C2. The C2 subset is small (n = 15) and has a higher proportion of females (66.7%) compared with C1 (44.1%) and the unassigned group (48.5%).

## Discussion

The present findings indicate that neuroanatomical dimensions of major depressive disorder (MDD), previously identified in the COORDINATE-MDD consortium and externally validated in the UK Biobank (Fu et al., 2024; Xiao et al., 2025), are detectable in the PRONIA cohort of young adults with recent-onset depression. Application of the pretrained HYDRA model without retraining identified two clusters corresponding to the previously described dimensions, supporting the reproducibility of this neuroanatomical framework in an independent multisite early-course sample. The present findings extend this framework by show that the clusters (C1, C2) can already be detected near illness onset, before the confounding effects of repeated depressive episodes and cumulative treatment exposure.

The detection of these dimensions in recent-onset depression supports the view that MDD heterogeneity reflects more than variation in symptom severity alone. The clusters did not differ significantly in overall depressive symptom severity, PANSS positive symptoms, or PANSS general psychopathology, but C1 showed higher negative symptom severity on both PANSS negative and SANS scores. This suggests that C1 may capture a dimension of early depression associated with motivational, expressive, and social withdrawal-related impairment rather than a simple gradient of depressive severity, consistent with MDD not as a unitary clinical syndrome but as a heterogeneous condition in which different symptom domains may map onto partially distinct neurobiological processes (Mayberg, 2003; Williams, 2017; Fu et al., 2019).

The presence of C1 and C2 expression in both healthy controls and individuals with recent-onset depression indicates that these patterns do not map directly onto categorical diagnoses, but instead reflect dimensions of neuroanatomical variation, consistent with the HYDRA framework (Varol et al., 2017). In PRONIA, C1 contained proportionally more individuals with recent-onset depression, whereas C2 showed a more balanced distribution of recent-onset depression and healthy control participants. This suggests that the dimensions may index neurobiological vulnerability that becomes clinically relevant when they interact with environmental factors, consistent with transdiagnostic brain-inflammation phenotypes observed across recent-onset depression and psychosis in PRONIA (Lalousis et al., 2022).

In peripheral inflammatory markers, IL-2 was significantly elevated in C1 with trends for IL-1β and IL-4, indicating an immune signal in the structurally preserved cluster. This pattern aligns with the inflammatory profile which differentiated recent-onset depression from healthy controls in PRONIA (Popovic et al., 2026), in which IL-1β, IL-2, IL-4 and related markers distinguished recent-onset depression from healthy controls. The present findings show that this inflammatory profile is predominant in C1 rather than uniformly distributed. IL-2 has a central role in T-cell proliferation, differentiation and immune homeostasis (Boyman and Sprent, 2012), and is essential for the development, survival, and suppressive function of regulatory T cells (Tregs), which constrain peripheral inflammation. Hypofunction of the Treg compartment has been proposed as a transdiagnostic mechanism contributing to the inflammatory phenotypes observed in depression and psychosis (Corsi-Zuelli and Deakin, 2021), and low-dose IL-2 has been shown to expand circulating Tregs and to potentiate antidepressant response in unipolar and bipolar depression (Foley et al., 2024), with a phase 2 randomised placebo-controlled trial in bipolar depression confirming Treg expansion alongside symptomatic improvement (Leboyer et al., 2025). Elevated peripheral IL-2 has previously been reported in MDD more broadly, where it may reflect active immune engagement (Dahl et al., 2014). The C1-localised elevation of IL-2 observed here may thus reflect a compensatory immunoregulatory response at illness onset rather than a systemic pro-inflammatory state. This is consistent with the more favourable response to SSRI antidepressants observed in D1 in COORDINATE-MDD (Fu et al., 2024), and suggests that a preserved capacity for Treg-mediated immune regulation may be one biological correlate of standard antidepressant responsiveness in early depression.

The inflammatory profile observed in C1 contrasts with our previous UK Biobank finding, in which D2 was associated with a broader pro-atherogenic and inflammatory metabolic profile alongside cognitive impairment, childhood adversity, self-harm, and distinct genetic loci, in an older sample with longer illness duration (Xiao et al., 2025). In the present recent-onset cohort, inflammation was most evident in the structurally preserved C1 cluster, indexed by elevated IL-2, whereas the structurally impaired C2 cluster was characterised by higher residual proteomic age. Peripheral cytokine alterations and proteomic ageing appear to be partially dissociable rather than co-localised within the same neuroanatomical dimension at illness onset.

The association of C2 with higher residual proteomic age is consistent with prior COORDINATE-MDD work in which D2 is associated with a greater MRI-derived brain age gap, with effects most pronounced at older chronological ages (Sharma et al., 2026). Together, the findings suggest that D2 may index a dimension shaped by cumulative environmental and biological exposure, such as chronic stress, early-life adversity, metabolic dysregulation, and chronic low-grade inflammation, which is superimposed on the genetic susceptibility identified in UK Biobank (Xiao et al., 2025). C1 may capture a more endogenous neurobiological phenotype, with a specific cytokine signal already detectable at illness onset and a more favourable response to SSRI antidepressants relative to placebo (Fu et al., 2024).

A mechanistic framework linking environmental adversity, accelerated biological ageing, and chronic inflammation provides a plausible substrate for the C2/D2 phenotype. Cumulative environmental exposures dysregulate the hypothalamic-pituitary-adrenal axis and sympathetic nervous system, induce glucocorticoid receptor desensitisation, and engage the Conserved Transcriptional Response to Adversity, which upregulates pro-inflammatory gene expression in circulating leukocytes (Cole, 2019). Sustained low-grade inflammation in turn induces cellular senescence and the senescence-associated secretory phenotype, generating a feed-forward inflammatory loop (“inflammaging”) that accelerates biological ageing across multiple organ systems (Franceschi et al., 2018; Furman et al., 2019). The proteomic age clock in the present study captures this convergence as the proteins contributing most strongly to its chronological-age prediction are involved in immune response and inflammation, alongside extracellular matrix remodelling and hormone regulation (Argentieri et al., 2024). In parallel, hgh-dimensional immune profiling has demonstrated that an immune-cell composite (“IMM-AGE”) predicts all-cause mortality independently of chronological age, supporting a quantifiable trajectory of immune ageing relevant to age-related disease risk (Alpert et al., 2019). Environmental exposures further explain substantially more variance in proteomic age and mortality than polygenic risk scores (Argentieri et al., 2025), and accelerated epigenetic and proteomic ageing has been shown to mediate part of the association between adverse childhood experiences and depressive symptoms (Klopack et al., 2022). Within this framework, the higher residual proteomic age observed in C2 is consistent with a dimension of depression in which cumulative environmental burden may drive an integrated immune-senescence phenotype that becomes increasingly pronounced with longer illness duration and chronological age.

The present findings extend the previous transdiagnostic stratification of PRONIA, in which HYDRA trained on combined recent-onset depression and psychosis (Lalousis et al., 2022) identified a preserved and an impaired cluster with broadly comparable clinical structure. In the transdiagnostic analysis, the impaired cluster showed elevated CRP and tumour necrosis factor α in multivariate biomarker comparisons, and chronic-illness samples were preferentially assigned to the impaired cluster on external validation, consistent with a chronicity gradient. The present study has applied an externally pre-trained MDD-specific HYDRA model, which demonstrates the preserved (C1)/impaired (C2) structure in recent-onset depression itself and identifies an inflammatory signal at illness onset specifically within the preserved C1 cluster (IL-2). Together with the broader pro-atherogenic, inflammatory metabolic profile observed in D2 in the older UK Biobank sample (Xiao et al., 2025), these observations are suggest a stage-dependent shift in the dimensional localisation of inflammation from a specific, plausibly compensatory IL-2 signal in the preserved cluster (C1) at illness onset, through broader peripheral inflammation (CRP, TNF-α) in the transdiagnostic impaired cluster (C2), toward the full immuno-metabolic and structural phenotype that characterises D2 in chronic and population-level samples.

Limitations of the present study include the cross-sectional analysis, which precludes inferences about the temporal stability of the clusters or their relationship to longitudinal clinical outcomes, such as symptom progression or treatment response. The proteomic analyses were conducted in a relatively small subset, limiting power. A substantial proportion of participants were not assigned to either dimension, which may reflect intermediate or heterogeneous profiles.

In summary, neuroanatomically defined MDD dimensions are present early in the course of illness and are associated with distinct clinical profiles and biological signals in PRONIA cohort. These findings support the COORDINATE-MDD dimensional, biologically informed framework and highlight the potential for stratified approaches to improve treatment selection. The findings support a stratified treatment paradigm in MDD, in which biomarker-based subtyping complements clinical assessment to guide personalised care.

## Data Availability

Data supporting the findings of this study are available from the corresponding authors upon reasonable request, subject to institutional approvals, data sharing agreements, and applicable ethical and legal restrictions.

